# How attaining the goals of a stroke care pathway is related to functional outcome after stroke. A nationwide study

**DOI:** 10.1101/2023.03.16.23287389

**Authors:** Elin Bergh, Torunn Askim, Ole Morten Rønning, Hild Fjærtoft, Bente Thommessen

**Author notes:** Corresponding author:* Elin Bergh, MD, Department of Neurology, PO Box 1000, 1478 Lørenskog, Norway.

## Abstract

**Background and purpose:** In 2018 a stroke care pathway (SCP) was introduced in Norway. The goal of the pathway was to improve acute stroke care by reducing time-delay. We aimed to evaluate if achieving the target times of the SCP was associated with a better functional outcome at 3 months post-stroke and to identify characteristics of patients attaining the goals.

**Methods:** We performed a register-based study with data from the Norwegian stroke register (NSR). Patients registered with acute stroke in 2019 were included. Functional outcome at 3 months in the patients with “achievement” was compared to patients with “non-achievement”. Achievement was defined as attaining three important goals in the SCP; 1) time from symptom onset to contact with the emergency medical service, 2) time from symptom onset to arrival in hospital, 3) time from arrival in hospital to admission to a stroke unit. Additionally, we evaluated time from arrival in hospital to treatment with intravenous thrombolysis. Modified Rankin Scale (mRS) was used to measure functional outcome at 3 months post-stroke and functional independence was defined as mRS 0-2. Characteristics of the “achievers” were analysed by univariate and multivariate logistic regression analyses.

**Results:** In total 2818 patients were included, 460 (16%) were in the achievement group. The mean (SD) age was 72.5 (12.6) years and 1201 (43%) were women. The probability of being independent at 3 months post-stroke was significantly higher in the achievement group versus the non-achievement group (OR 1.36, 95% CI 1.04-1.79, p=0.026). The “achievers” were significantly younger, less likely to be living alone, less likely to have diabetes, more often had an ischemic stroke and were admitted with more severe strokes than the “non-achievers”.

**Conclusion:** Goal achievement in the SCP was significantly associated with independence 3 months post-stroke.

## Introduction

A care pathway aims to standardize and optimize patient care based on the best available evidence and guidelines for a specific condition.^1^ The intention of their implementation is to provide up-to-date diagnostics and treatment, to improve patient’s outcome and increase the cost effectiveness. Yet, their impact on patient outcome is under debate. ^2, 3^

It is well known that the results of revascularization therapies are better with earlier treatment and some studies demonstrate that early arrival in hospital is associated with a favourable outcome even when the patients do not receive reperfusion treatment. ^4, 5^ In addition, treatment in a stroke unit with a multidisciplinary team makes patients more likely to survive and increase their probability of being independent and living at home one year after stroke.^6^ Some studies indicate that functional outcome is improved by early admission to and more of the in-hospital time spent in a stroke unit. ^5, 7, 8^ All of this demonstrates that it is important to reduce pre- and intrahospital delay and that early admission to hospital is associated with better functional outcome.

Based on this knowledge, a national standardized pathway of stroke care (SCP) was established by the Norwegian Health authorities in 2017. The aim of this pathway was to ensure best stroke care and equal treatment throughout the country by reducing delay in assessment, diagnostics, and treatment of acute stroke. The SCP was introduced in 2018. ^9^ It is organized in two different phases and in the present study, we evaluate the first phase that covers the period from onset of acute stroke until discharge from hospital. The SCP includes 10 recommended target times, and the goal is to ensure achievement of the target-times in up to 80% of the patients. See table 1.

**Table 1.** The standardized pathway of stroke care; Points of measurement, recommended time-use and goals. The yellow colour indicates the goals that are analyzed in our study.

| Point of measurement | Recommended time-use | Goal |
| --- | --- | --- |
| **1. Time for onset of symptoms** |  | Registration only |
| **2A. Time from onset to contact with the emergency medical service (EMS)** | EMS should be contacted within 15 minutes from onset of symptoms. | 50 % of patients with AIS should contact EMS within 15 minutes |
| **2B. Time from contact with ambulance to arrival of ambulance** | The ambulance should be on place within 25 minutes from the call was made. | In 80 % of cases the ambulance should arrive within 25 minutes from the call was made. |
| **2C. Time from arrival to departure of ambulance.** | The EMS should leave within 25 minutes after arriving by the patient. | 80 % of the patients with symptoms of AIS should be transported to hospital within 25 minutes of arrival of the EMS. |
| **3. Time for hospitalization** | Time from onset of symptoms until arriving hospital. | 60 % of the patients should arrive hospital within 4 hours from onset of symptoms. |
| **4. Time for radiological examination** | Time from arriving hospital until CT/MRI is performed. | 60 % of the patients arriving within 4hours should receive CT/MR within 15 minutes. |
| **5. Time for thrombolysis** | Time from arrival at hospital until start of intravenous thrombolysis should be <40 minutes. | 60 % of patients receiving thrombolysis should start treatment within 40 minutes upon arrival. |
| **6A. Thrombectomy** | Time from onset of symptoms until the patient is hospitalized in a center performing thrombectomy. | Only registration. |
| **6B. Thrombectomy** | Time from hospitalization until arterial puncture is performed. | Patients directly admitted to an intervention-centre: 50 % should be punctured within 60 minutes.<br>Patients transferred from another hospital: 50 % should be punctured within 40 minutes |
| **6C. Thrombectomy: Time for recanalization, TICI 2 or 3.** | Time from start of thrombectomy until recanalization. | Only registration. |
| **7. Time to admission to a stroke unit.** | Time from arrival in hospital to admission to a stroke unit should be <3 hours. | 80 % of patients with acute stroke should be admitted to a stroke unit within 3hours. |
| **8A. Time to examination of a possible stenosis of arteria carotis interna (ICA).** | Time from admission to hospital until the examination should not exceed 3 days. | 60 % of patients with acute ischemic stroke should be examined for ICA stenoses within 3 days upon admission. |
| **8B. Time to carotid endarterectomy** | Time from onset of symptoms until carotid endarterectomy should be <14 days. | 80% of patients operated for a symptomatic ICA stenoses should be endarterectomized within 14 days. |
| **9. Interdisciplinary assessment in a stroke unit.** | Time from admission to a stroke unit until an interdisciplinary assessment is performed. | 80 % of the patients should receive interdisciplinary evaluation within 7 days of admission and always upon discharge. |
| **10 Time from the patients is ready to be discharged until he/she receives appropriate rehabilitation.** | Time from the patients is ready to be discharged until he/she receives rehabilitation. | Only registration. |

EMS = emergency medical service, CT = computed tomography, MRI = magnetic resonance image, ICA = internal carotid artery, AIS = acute ischemic stroke

In a previous study, we evaluated the impact of the pathway on functional outcomes after stroke. No change in global function measured by the modified Rankin Scale (mRS) or mortality 3 months post-stroke was found. However, more patients were treated in a dedicated stroke unit and more patients were discharged directly home from the acute hospital stay, compared to before the SCP was introduced.^10^

In the present study we have chosen to focus on three essential goals in the pathway related to rapid admission to hospital and stroke unit to assess goal achievement.

Our primary hypothesis was that patients who achieved all three target times would have significantly better functional outcome at 3 months post-stroke compared to those experiencing a delay.

## Methods

### Study design, setting and population

The present study was a prospective study using data obtained in the acute phase and at 3 month follow up from the Norwegian Stroke Register (NSR). The NSR is a compulsory register where all Norwegian acute care hospitals are obliged to report medical data on patients ⩾18 years admitted to hospital with acute stroke. It is estimated that the register has a coverage of 87% of all patients with acute stroke. In Norway stroke diagnostics and treatment is provided by public healthcare service that is publicly funded and equal to all residents. Norwegian stroke care guidelines state that all patients with the diagnosis of acute stroke after management in the emergency department, should rapidly be transferred to a multidisciplinary stroke unit for further examinations and treatment. ^11^ In 2019 the population of Norway was 5.3 million and about 10000 patients were diagnosed with acute stroke, of which 9000 patients were registered in the NSR. Data for 10 recommended goals in the stroke care pathway were incorporated in the NSR in 2018.

All patients registered in the NSR with acute stroke in 2019 and with a follow up assessment 3-months post-stroke were included in the present study. Figure 1 shows the study population that met the inclusion criteria.

**Figure 1.**
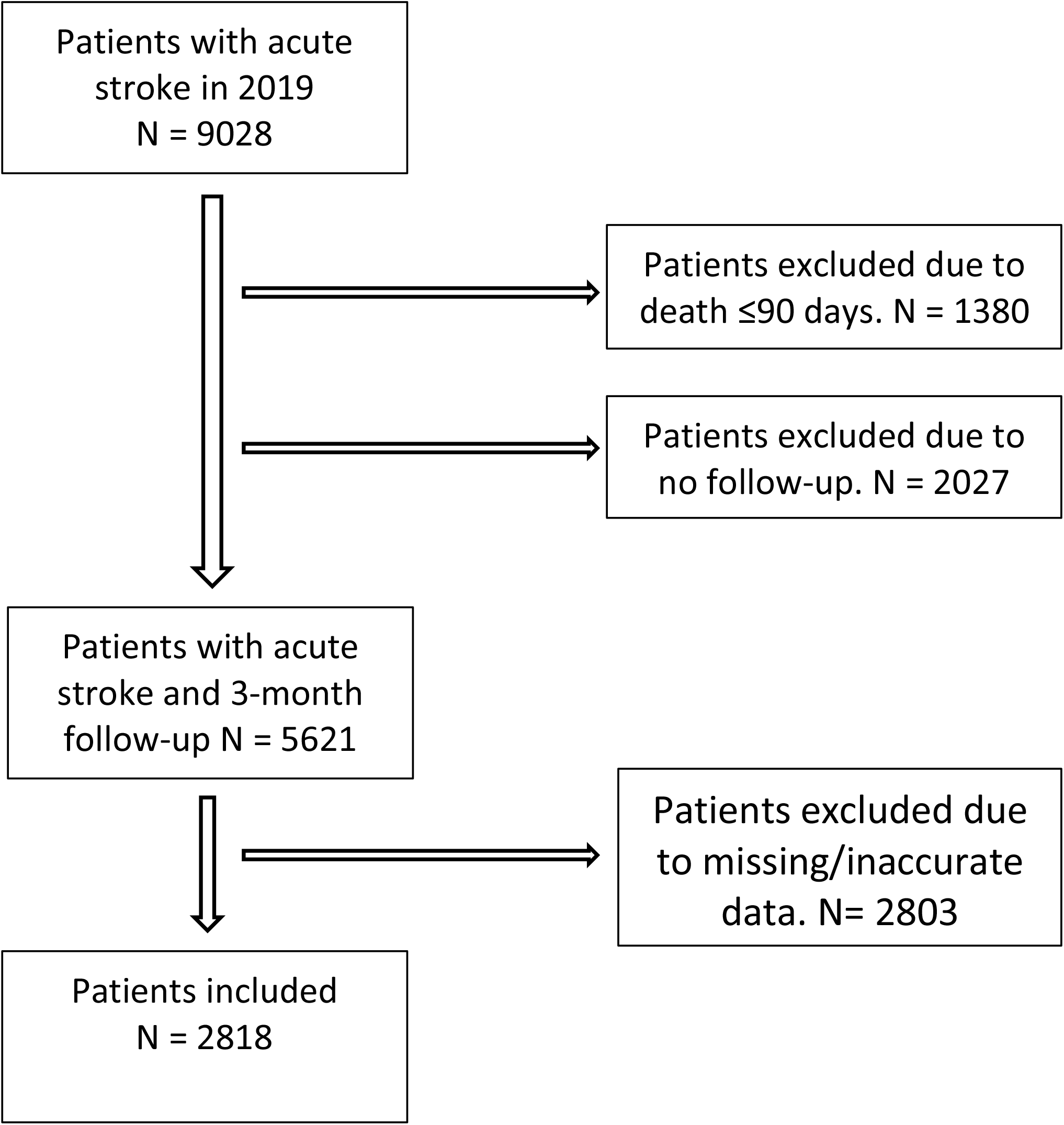
Flow chart showing inclusion/exclusion of patients.

The first phase of the stroke care pathway consists of 10 recommended goals for time-use from onset of symptoms until the patient is ready to be discharged from hospital as shown in table 1. In the present study, we have chosen three essential goals in the pathway based upon their importance for functional outcome and that sufficient and quantifiable data are present to evaluate goal achievement.

1. Time from symptom onset to contact with the emergency medical service (EMS) ≤ 15 minutes (goal #2A).
2. Time from symptom onset to admission to hospital ≤ 4 hours (goal #3).
3. Time from arrival in hospital to admission to a stroke unit ≤ 3 hours (goal #7).

“Achievement” was defined as fulfilling all 3 goals within the recommended target times.

“Non-achievement” was defined as fulfilling 0 - 2 of these goals.

In addition to these three goals, time from hospital admittance to thrombolysis less than 40 minutes was included in a sub analysis (goal #5).

### Data collection

The data retrieved from the Norwegian Stroke Register comprise:

1. Baseline characteristics: age, sex, marital status, residence, health region.
2. Cardiovascular risk factors: hypertension, atrial fibrillation, previous transient ischemic attack (TIA), previous stroke, diabetes mellitus.
3. Functional status: neurological deficit at admission measured by the National Institutes of Health Stroke Scale (NIHSS), functional status before and three months post-stroke measured by modified Rankin Scale. ^12 13^
4. Time points in the stroke care pathway: time for onset of symptoms, time for contact with the EMS, time for hospitalization, time for thrombolysis, time for admission to a stroke unit.

### Outcomes

Primary outcome was the probability of good functional outcome (mRS 0-2) 3 months post-stroke in the achievement group compared to patients in the non-achievement group. mRS is a 6-point disability scale ranging from 0 to 5 where 0 corresponds to no symptoms and 5 corresponds to severe disability. A score of 6 denotes death. The modified Rankin Scale is a widely used outcome measure in stroke trials and multiple studies have demonstrated its validity and reliability. ^14^ Evaluation of the mRS before the stroke was based on information from the patient, their next of kin and the patients’ medical record. mRS at 3 months was assessed by health personnel at a 3 month follow up assessment either by phone or at the outpatient clinic.

Secondary outcomes were characteristics of patients in the achievement group and variation between the different health regions.

### Statistics

Comparison of mRS (0-2) at 3 months post-stroke in the achievement versus non-achievement group was defined as the primary analysis. Logistic regression analyses were applied using mRS 0-2 (yes vs. no) as the outcome variable and achievement (yes vs. no) as the independent variable and adjusting for age, sex and pre-stroke mRS (0-2 vs. 3-5).

Descriptive data were presented as median and interquartile range (IQR) for categorical variables and means and standard deviations (SDs) for continuous variables. The patients in the achievement group were compared to patients in the non-achievement group using the *t*-test Wilcoxon signed-rank test (continuous data) or the *χ*^2^ test (categorical data).

Logistic regression analyses for achievement of the pathway goals (yes or no, as the dependent variable) were used to estimate the influence of predefined characteristics. In the univariate logistic regression analyses age, sex, living alone, hypertension, diabetes mellitus, prior TIA, prior stroke, atrial fibrillation, etiology of stroke (ischemic or. hemorrhagic), stroke severity (NIHSS) at admission, pre-stroke mRS (0-2 or 3-5) and health region were used as independent variables. Outcome is presented as Odds Ratios (OR) with 95% CIs for the main results.

Independent variables with p≤0.05 in the univariate analyses were subjected into the multivariate logistic regression analysis.

Significance levels were set at p≤ 0.05, using two-sided test.

Statistical analyses were conducted using SPSS version 27 (IBM SPSS, Chicago, IL, USA).

## Ethics

The Regional Committee for Medical Research Ethics Central Norway (REC no 80125). approved the study. In accordance with the approval from the Regional Committee for Medical Research Ethics and the Norwegian law on medical research, the project did not require a written patient consent.

## Results

In total 9028 patients with acute stroke were registered in NSR in 2019. Of these, 1380 (15%) patients died within 3 months and 2027 (23%) were excluded due to no 3-month follow-up assessment. In addition, 2803 (31%) were excluded due to missing or inaccurate data. The missing data were mostly due to missing registration of time for contact with the EMS. Altogether 2818 (31%) of all patients registered in 2019 were included in the present study. See figure 1.

In total 460 (16%) of the 2818 patients achieved all 3 goals. Number of patients achieving the goals are displayed in table 2. The characteristics of patients achieving the goals versus the non-achievers are compared in table 3. The patients in the achievement group were significantly younger, were not living alone, were non-diabetics and had more often a hemorrhagic stroke. The stroke severity (NIHSS) at admission was significantly higher in the achievement group, 6.0 versus 4.7.

**Table 2.**
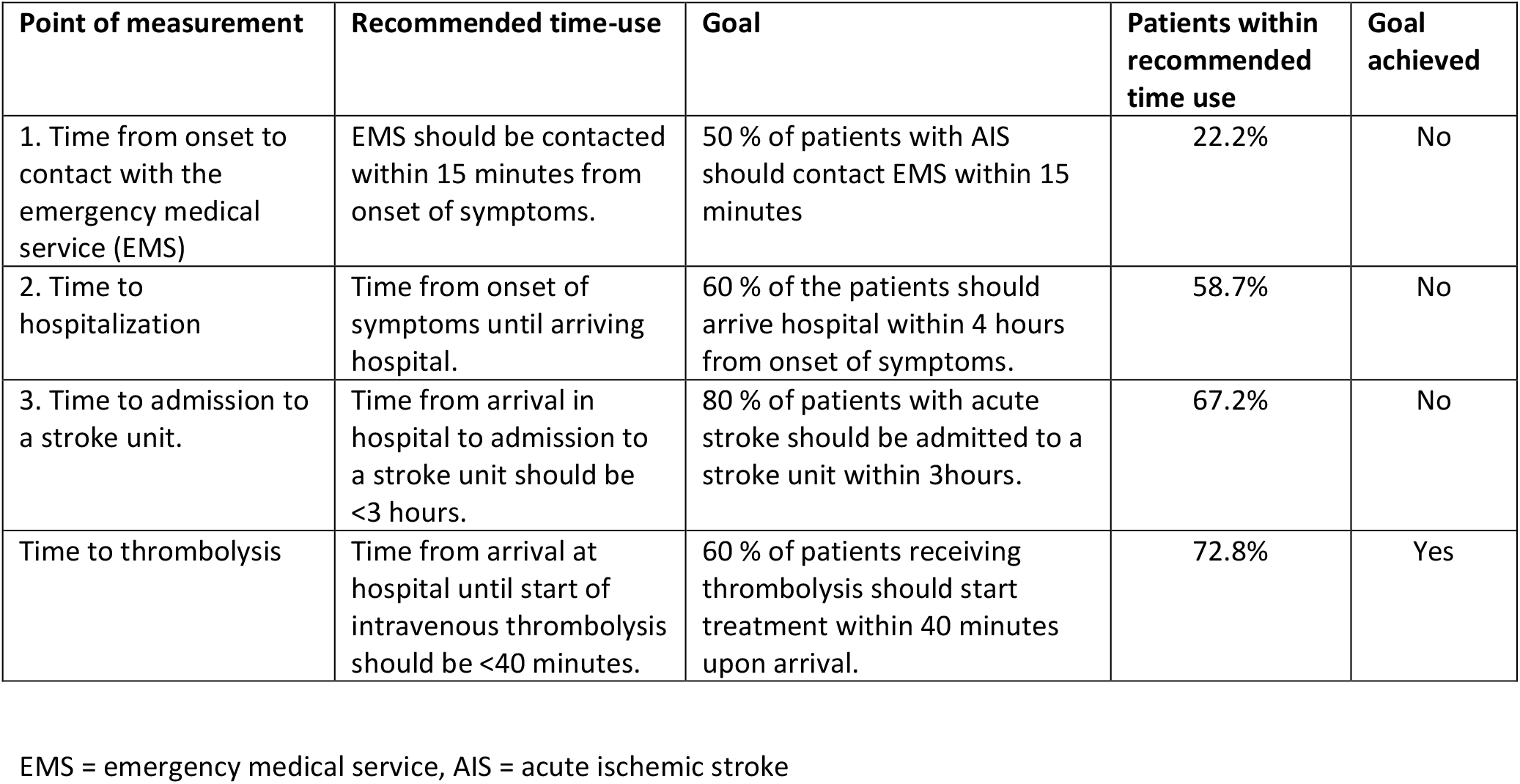
The standardized pathway of stroke care with proportion of patients achieving recommended goals.

| Point of measurement | Recommended time-use | Goal | Patients within recommended time use | Goal achieved |
| --- | --- | --- | --- | --- |
| 1. Time from onset to contact with the emergency medical service (EMS) | EMS should be contacted within 15 minutes from onset of symptoms. | 50 % of patients with AIS should contact EMS within 15 minutes | 22.2% | No |
| 2. Time to hospitalization | Time from onset of symptoms until arriving hospital. | 60 % of the patients should arrive hospital within 4 hours from onset of symptoms. | 58.7% | No |
| 3. Time to admission to a stroke unit. | Time from arrival in hospital to admission to a stroke unit should be <3 hours. | 80 % of patients with acute stroke should be admitted to a stroke unit within 3hours. | 67.2% | No |
| Time to thrombolysis | Time from arrival at hospital until start of intravenous thrombolysis should be <40 minutes. | 60 % of patients receiving thrombolysis should start treatment within 40 minutes upon arrival. | 72.8% | Yes |

EMS = emergency medical service, AIS = acute ischemic stroke

**Table 3.** Patient characteristics in total and stratified by achievement of the goals in the stroke care pathway.

|  | Total<br>N = 2818 | Achievement<br>group<br>N = 460 (16.3%) | Non-achievement<br>group<br>N = 2358 (83.7%) | P- value |
| --- | --- | --- | --- | --- |
| **Demographics** |  |  |  |  |
| Age, mean (SD), y | 72.5 (12.6) | 71.6 | 73.7 | 0.001 |
| Female sex, n (%) | 1201 (42.6) | 189 (41.6) | 1012 (42.9) | 0.47 |
| Living alone pre-stroke | 1076 (38.2) | 131 (28.5) | 945 (40.1) | <0.001 |
| **Risk factors** |  |  |  |  |
| Hypertension | 1647 (58.4) | 260 (56.5) | 1387 (58.8) | 0.36 |
| Diabetes Mellitus | 497 (28.2) | 58 (12.6) | 439 (18.6) | 0.002 |
| Previous TIA | 277 (9.8) | 44 (9.6) | 233 (9.9) | 0.77 |
| Previous stroke | 661 (23.5) | 95 (20.7) | 566 (24.0) | 0.12 |
| Previous atrial fibrillation | 646 (22.9) | 99 (15.3) | 547 (23.2) | 0.44 |
| **Ischemic stroke** | 2535 (90.0) | 400 (87.1) | 2135 (91.0) | 0.009 |
| **Degree of disability** |  |  |  |  |
| NIHSS at admission, mean, |  | 6.0 | 4.7 | <0.001 |
| mRS pre-stroke, n (%) |  |  |  |  |
| - mRS 0-2 | 2331 (82.7) | 399 (92.8) | 1932 (88) | 0.004 |
| - mRS 3-5 | 294 (10.4) | 31 (6.2) | 263 (12) |  |

TIA – transient ischemic attack
\*Each region vs. the other 3 regions

At 3 months post-stroke the probability of having good functional outcome was significantly higher in the achievement group versus the non-achievement group (OR 1.36, 95% CI 1.04-1.79, p = 0.026) after adjusting for age and sex and pre-stroke mRS.

The multivariable logistic regression model showed that achieving the goals of the pathway was associated with living with someone pre-stroke (OR 1.54, 95% CI 1.20-1.98, p= 0.001), and with more severe stroke at admittance to hospital (OR 1.46, 95% CI1.15-1.85, p=0.002) and negatively associated with diabetes mellitus (OR 1.48, 95% CI1.06-2.07, p =0.02) and a trend towards being dependent pre-stroke (OR 0.65, 95% CI 0.42-0.99, p=0.05). See table 4.

**Table 4.** Univariate and multivariable logistic regression analyses. Characteristics of patients achieving the goals of the stroke care pathway.

|  | N = 2818 | Bivariate |  |  | N = 2094 | Multivariable |  |  |
| --- | --- | --- | --- | --- | --- | --- | --- | --- |
|  |  | Odds ratio | 95% CI | p-value |  | Odds ratio | 95% CI | p-value |
| Age | 2818 | 0.99 | 0.98 – 0.99 | **0.001** | 2094 | 0.99 | 0.99 – 1.00 | 0.21 |
| Sex |  |  |  |  |  |  |  |  |
| Men | 1617 | 1.00 |  |  | 1174 | 1.00 |  |  |
| Women | 1201 | 0.93 | 0.76 – 1.14 | 0.47 | 920 | 1.04 | 0.82 – 1.32 | 0.76 |
| Ischemic stroke |  |  |  |  |  |  |  |  |
| Yes | 2535 | 1.00 |  |  | 1897 | 1.00 |  |  |
| No | 269 | 1.50 | 1.10 – 2.04 | **0.01** | 197 | 1.33 | 0.92 – 1.91 | 0.13 |
| Living alone pre-stroke |  |  |  |  |  |  |  |  |
| Yes | 1076 | 1.00 |  |  | 805 | 1.00 |  |  |
| No | 1727 | 1.67 | 1.34 – 2.08 | **<0.001** | 1289 | 1.54 | 1.20 – 1.98 | **0.001** |
| Health region |  |  |  |  |  |  |  |  |
| 1 | 1238 | 1.00 |  |  | 894 | 1.00 |  |  |
| 2 | 679 | 0.60 | 0.46 – 0.79 | **<0.01** | 533 | 0.58 | 0.43 – 0.80 | **0.001** |
| 3 | 589 | 0.99 | 0.77 – 1.28 | 0.93 | 446 |  |  |  |
| 4 | 312 | 0.84 | 0.60 – 1.18 | 0.31 | 221 |  |  |  |
| Diabetes mellitus |  |  |  |  |  |  |  |  |
| Yes | 497 | 1.00 |  |  | 359 | 1.00 |  |  |
| No | 2317 | 1.58 | 1.18 – 2.13 | **0.002** | 1735 | 1.48 | 1.06 – 2.07 | **0.02** |
| Anti-hypertensive treatment |  |  |  |  |  |  |  |  |
| Yes | 1647 | 1.00 |  |  |  |  |  |  |
| No | 1165 | 1.10 | 0.90 – 1.35 | 0.36 |  |  |  |  |
| Previous TIA |  |  |  |  |  |  |  |  |
| Yes | 277 | 1.00 |  |  |  |  |  |  |
| No | 2498 | 1.05 | 0.75 – 1.48 | 0.77 |  |  |  |  |
| Previous stroke |  |  |  |  |  |  |  |  |
| Yes | 661 | 1.00 |  |  |  |  |  |  |
| No | 2144 | 1.21 | 0.95 – 1.55 | 0.12 |  |  |  |  |
| Atrial fibrillation |  |  |  |  |  |  |  |  |
| Yes | 646 | 1.00 |  |  |  |  |  |  |
| No | 2162 | 1.10 | 0.86 – 1.40 | 0.44 |  |  |  |  |
| MRS pre-stroke |  |  |  |  |  |  |  |  |
| mRS 0 - 2 | 2331 | 1.00 |  |  | 1844 | 1.00 |  |  |
| mRS 3-5 | 294 | 0.57 | 0.39 – 0.84 | **0.005** | 250 | 0.65 | 0.42 – 0.99 | **0.05** |
| Stroke severity at admission |  |  |  |  |  |  |  |  |
| NIHSS 0-5 | 1497 | 1.00 |  |  | 1397 | 1.00 |  |  |
| NIHSS > 5 | 748 | 1.50 | 1.20 – 1.88 | **<0.001** | 697 | 1.46 | 1.15 – 1.85 | **0.002** |

TIA – transient ischemic attack, mRS – modified Rankin Scale, NIHSS – National Institutes of Health Stroke Scale

Significant regional differences were found and living in one of the regions was identified to be negatively associated with goal achievement both in the main and the subgroup analyses.

By analysing the characteristics of patients receiving thrombolysis with a door-to-needle time ≤ 40 minutes, we found that not suffering from atrial fibrillation (AF) was positively associated with receiving thrombolysis within the time limit. See table 5

**Table 5.**
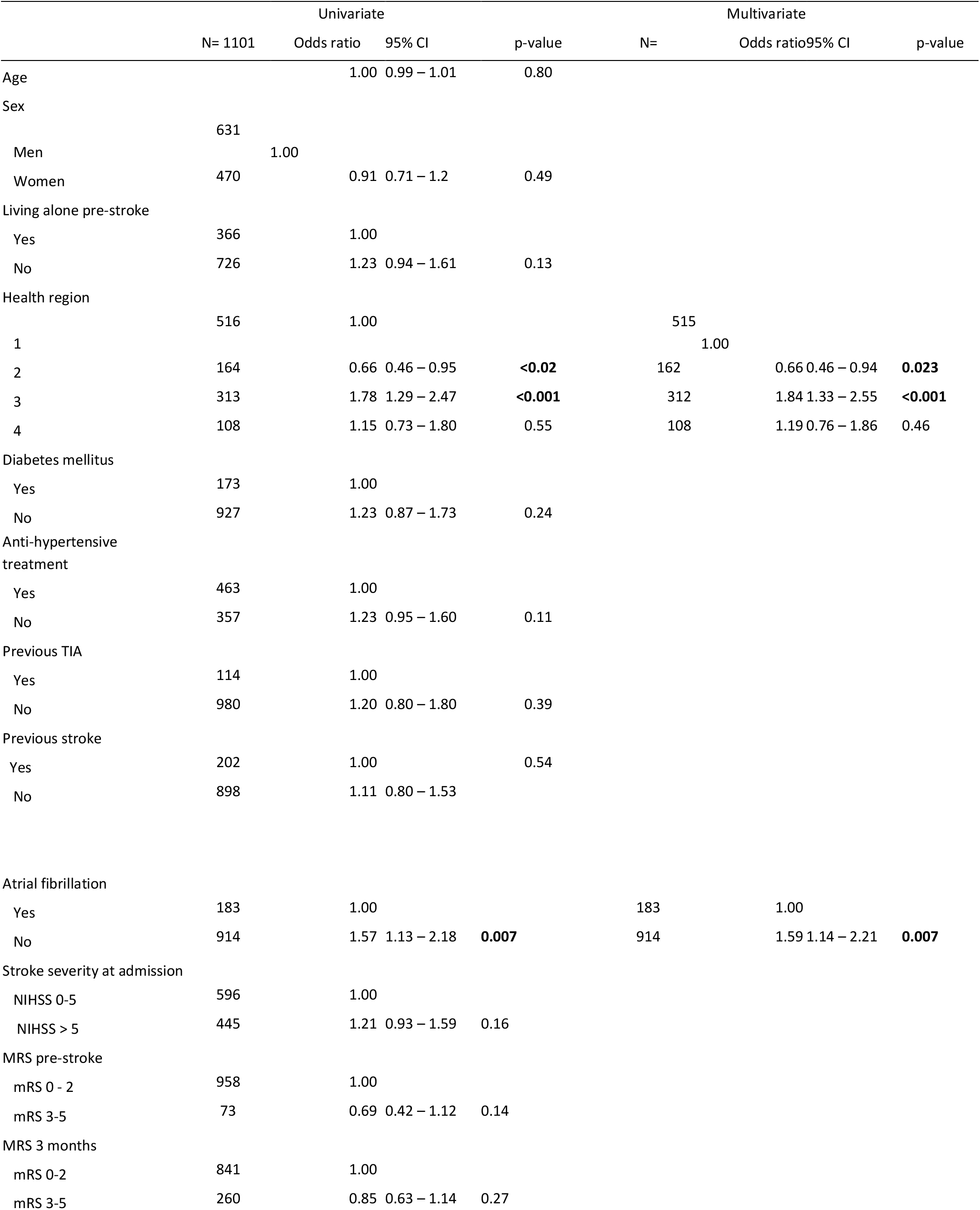
Univariate and multivariable logistic regression analyses. Predictors for time from hospitalization to thrombolysis <= 40min.

## Discussion

The main finding in our study was that the odds of having good functional outcome 3 months after acute stroke was significantly better in the group of patients achieving the chosen goals of the stroke care pathway compared to the group of non-achievers. Living with someone, suffering from severe stroke, a trend towards being independent prior to the stroke and not having diabetes mellitus were the individual factors associated with good achievement of the target times.

The pathway focuses on reducing time delay, which is vital for a good treatment outcome in acute stroke. Early contact with the EMS and early admission to hospital are factors facilitating early revascularization treatment, which is highly time dependent. The sooner this treatment is given the better is the outcome at 3 months post-stroke. The odds ratio (OR) of an excellent outcome (being independent) at 3 months with thrombolysis compared with placebo decreases with treatment delays, from 2.8 when patients were treated between 0 and 90 min, to 1.2 for 271 to 360 min. ^15^

In our study only 16% of the patients achieved the three chosen target goals. Regarding the pre-hospital measure points a majority of the patients were admitted to hospital within 4 hours from symptom onset while only 22% of the patients were contacting the EMS within 15 minutes.

The main causes of pre-hospital delay may to some degree be explained by the challenge in recognizing stroke symptoms by the patients and their bystanders and sometimes by the EMS-personnel and the general practitioners as well. ^16^ A broad variation of symptoms and the possibility of impaired communication due to speech disturbances may also cause delay in recognition as may the distance to the EMS and the distance to hospital.^17 16^

Over the last decades several studies have looked at the pre-hospital delay in acute stroke and how it can be reduced.^16, 18^ Campaigns focusing on increasing stroke awareness in the public through mass media and social media are shown to be effective, but the effect declines after campaign closure. A Norwegian study from 2016 showed increased rate of patients admitted to the emergency room and receiving thrombolysis after such a campaign, but the effect tapered off after 3 to 6 months implying that these campaigns need to be repeated.^19^ The last stroke campaign in Norway prior to January 2019 took place from October 2016 to January 2017. ^20^

In addition to pre-hospital delay, intra-hospital factors like specific stroke treatment protocols, emergency department effectiveness, organization of SU and brain imaging room within the hospital are of importance regarding treatment delay. ^21^

Our results showed that only 67% of the patients were admitted to a stroke unit within three hours from admission to hospital. To reduce the time from arriving hospital to admission to the stroke unit, rapid and streamlined management in the emergency department is required. In addition, the capacity of the stroke unit to receive new patients is a prerequisite to reduce delayed admission to the stroke unit.

There are no accurate statistics regarding number of beds in a stroke unit in Norway. Data from the NSR from 2019 shows that 86% of the patients were directly admitted to a stroke unit while 94% of the patients were treated in a stroke unit during their hospital stay. Recognizing an acute stroke is not always straight forward. Some strokes may be misdiagnosed as another medical condition and hence may have a delayed transfer to the stroke unit. It is estimated that more than 30% of the patients admitted for stroke are stroke mimics. ^22, 23^ The capacity of stroke units to receive new patients may therefore be reduced by an increasing number of patients not having a stroke.

Characteristics of patients who achieved the goals of the SCP differed substantially from those with non-achievement, most notably by not living alone prior to the stroke. Patients attaining the goals were also more independent pre-stroke but had a more severe stroke at admission to hospital in accordance with previous studies. ^18, 24^

Living alone may be a disadvantage regarding early contact with the EMS and attending hospital as fast as possible is intuitive and is previously shown. ^25-27^ It is also reasonable and in line with earlier studies that patients with reduced functional status pre-stroke due to immobility or comorbidity are less able to contact someone when they experience symptoms of acute stroke. Additionally, symptoms of comorbidities might complicate the examination and make it more difficult to recognize symptoms as an acute stroke.^16^ In the present study diabetes was associated with delayed contact and admission. A systematic review of stroke in diabetics showed an inadequate knowledge of signs and symptoms of stroke.^28^ Stroke symptoms misinterpreted as symptoms of hypoglycemia may add to the increased risk of delay in diabetics. These findings indicate that patient characteristics influence stroke care pathway achievement and better attainment should probably in particular focus on reaching those living alone, with diabetes and with functional impairment.

Patients achieving the pathway goals had a more severe stroke at admission reflected by higher NIHSS compared to the non-achievement group. More severe stroke symptoms would possible have made it easier to recognize the symptoms as acute stroke, resulting in earlier contact with EMS and thereby reduced pre-hospital delay. This is in accordance with previous studies that have revealed that patients with severe strokes arrive hospital faster than patients with milder strokes.^18^

In addition to ensure that patients with acute stroke would receive well organized and predictable care without non-medical delay in assessment, diagnostics or treatment, the SCP intended to reduce variations in stroke care throughout the country. In Norway, stroke service was high functioning even before introduction of the SCP. Nevertheless, an important finding in our study was regional differences in time from onset of stroke to contact with EMS, to hospitalization, time from hospital admission to thrombolysis and to admission to a stroke unit.

In addition to significant regional differences, we found that not having atrial fibrillation was associated with receiving thrombolysis within 40 minutes. Possible explanations to why patients with atrial fibrillation receive tPA later could be insecurity whether they were on anticoagulation which might be a contraindication to tPA. Patients with atrial fibrillation are also older with more comorbidities.

Regional differences could originate in different organization of acute stroke treatment, different regional guidelines, local traditions/routines, distance to hospital and differences in population living in urban and rural settings. Patients living in rural areas have longer pre-hospital time intervals before treatment. ^17, 29^ Rapid and streamlined management in the emergency department to improve the in-hospital management are required, and this may unfortunately vary between the hospitals. According to the NSR, hospitals with more than 50 patients receiving thrombolytic treatment per year, are more likely to give tPA within 40 minutes.^22^

Some hospitals have focused on reducing time to thrombolysis for several years by routinely performing simulation-based team training. This means that all the personnel included in the “acute stroke team” practise the acute stroke procedure by simulating the admission of a real patient with acute stroke either by a simulation doll or patient. Observers watch the training and later discuss with the involved personnel what works well and what could be improved. In a study from Stavanger University Hospital, Norway, they found that the door-to-needle time was reduced by 13 minutes by simulation training and protocol revision.^30^ This training could be recommended for all hospitals with a stroke unit.

Implementation of a SCP and compulsory registration of target times in the NSR may help the individual hospital and the different health regions to compare their own stroke services with other hospitals in Norway, and to identify suboptimal organizational and treatment factors that need to be improved.

## Strengths and limitations

The large sample size and the high coverage rate of the NSR (87%) are strengths of the present study. Yet, using a register always includes a risk of inaccurate registration, incomplete and missing data. Excluding the patients without a 3-month follow-up and the patients with missing data left us with only 30% of the total patients.

This could result in a potential selection bias and reduce the generalizability of our study. In our previous study, we compared baseline characteristics of those included versus excluded, and found that the no follow-up cohorts were significantly older, had poorer functioning, and were institutionalized to a higher extent than the groups with a 3-month follow-up. ^10^

Due to missing and inaccurate data we have analysed only three out of 10 target goals of the pathway. However, the chosen goals, such as time from symptom onset to admission to hospital and thereby time to reperfusion therapy and time to admittance in a stroke unit are of significant importance regarding stroke outcome.

It is well documented that it takes time to change clinical practice and as our data were collected only 1 year after the SCP was introduced, uncertainty exists whether all hospitals had fully implemented the pathway. ^31^ Hence it might be too early to identify the effect of the pathway, especially regarding functional outcome. It is expected that further implementation of the SCP and attention about the regional differences revealed will increase the focus on reducing time delay.

It is also possible that improvement in target times is caused by a general change in clinical practice rather than implementation of the SCP, as national guidelines have emphasised fast treatment in the acute phase since 2010.

The mRS scale is used as a primary outcome measure. In mild to moderate stroke, the mRS seems to detect small and significant treatment effect changes. ^32^

## Conclusion

Although the basic structures of the stroke service in Norway were established prior to the introduction of the Stroke Care Pathway, our study shows that a favourable outcome is associated with achievement of three essential target times of the SCP. Living with someone was the most significant patient related factor associated with goal achievement. The regional differences indicate that the target times in the SCP may be used as a tool for improvement in stroke services.

## Data Availability

There are restrictions applied to the availability of the data referred to in this manuscript as they were used under license for this study.

